# Electrophysiological correlates of thalamocortical function in acute severe traumatic brain injury

**DOI:** 10.1101/2021.09.08.21262992

**Authors:** William H. Curley, Yelena G. Bodien, David W. Zhou, Mary M. Conte, Andrea S. Foulkes, Joseph T. Giacino, Jonathan D. Victor, Nicholas D. Schiff, Brian L. Edlow

## Abstract

Few reliable biomarkers of consciousness exist for patients with acute severe brain injury. Tools assaying the neural networks that modulate consciousness may allow for tracking of recovery. The mesocircuit model, and its instantiation as the ABCD framework, classifies resting-state EEG power spectral densities into categories reflecting widely separated levels of thalamocortical network function and correlates with outcome in post-cardiac arrest coma.

We applied the ABCD framework to acute severe traumatic brain injury and tested four hypotheses: 1) EEG channel-level ABCD classifications are spatially heterogeneous and temporally variable; 2) ABCD classifications improve longitudinally, commensurate with the degree of behavioural recovery; 3) ABCD classifications correlate with behavioural level of consciousness; and 4) the Coma Recovery Scale-Revised arousal facilitation protocol improves EEG dynamics along the ABCD scale. In this longitudinal cohort study, we enrolled 20 patients with acute severe traumatic brain injury requiring intensive care and 16 healthy controls. Through visual inspection, channel-level spectra from resting-state EEG were classified based on spectral peaks within frequency bands defined by the ABCD framework: ‘A’ = no peaks above delta (<4 Hz) range (complete thalamocortical disruption); ‘B’ = theta (4-8 Hz) peak (severe thalamocortical disruption); ‘C’ = theta and beta (13-24 Hz) peaks (moderate thalamocortical disruption); or ‘D’ = alpha (8-13 Hz) and beta peaks (normal thalamocortical function). We assessed behavioural level of consciousness with the Coma Recovery Scale-Revised or neurological examination and, in 12 patients, performed repeat EEG and behavioural assessments at ≥6-months post-injury.

Acutely, 95% of patients demonstrated ‘D’ signals in at least one channel but exhibited heterogeneity in the proportion of different channel-level ABCD classifications (mean percent ‘D’ signals: 37%, range: 0-90%). By contrast, healthy participants and patients at follow-up predominantly demonstrated signals corresponding to intact thalamocortical network function (mean percent ‘D’ signals: 94%). In patients studied acutely, ABCD classifications improved after the Coma Recovery Scale-Revised arousal facilitation protocol (*P*<0.05), providing electrophysiological evidence for the effectiveness of this commonly performed technique. ABCD classification did not correspond with behavioural level of consciousness acutely, where patients demonstrated substantial within-session temporal variability in ABCD classifications. However, ABCD classification distinguished patients with and without command-following in the subacute-to-chronic phase of recovery (*P*<0.01). Patients also demonstrated significant longitudinal improvement in EEG dynamics along the ABCD scale (median change in ‘D’ signals: 37%, *P*<0.05).

These findings support the use of the ABCD framework to characterize channel-level EEG dynamics and track fluctuations in functional thalamocortical network integrity in spatial detail.

## Introduction

Clinical evaluation of level of consciousness in patients with acute severe brain injury is limited by a lack of reliable biomarkers. The ‘mesocircuit’ model proposes that the central thalamus and its projections to the cerebral cortex are critical to supporting the recovery of consciousness.^1,2^ Disruptions in this thalamocortical network have been implicated in disorders of consciousness (DoCs) following severe brain injury.^3-6^ Tools that assay thalamocortical network integrity may therefore allow for more precise tracking of neurological recovery than is possible with behavioural assessments alone.

The mesocircuit model provides a framework for classifying power spectra of clinical, resting-state EEG into four categories (‘A,’ ‘B,’ ‘C,’ ‘D’). These categories correspond to widely separated levels of functional thalamocortical network integrity,^2^ and application of this ‘ABCD framework’ has been shown to correlate with outcome in patients with post-cardiac arrest coma.^7^ The ABCD categories and their functional correlates in the mesocircuit model are as follows. The ‘A’ category (no spectral peak above the delta [<4 Hz] range) corresponds to a completely disconnected thalamocortical network that can be likened to the ‘cortical slab preparation,’ wherein neocortical neurons are markedly hyperpolarized and exclusively produce low frequency oscillations.^8^ The ‘B’ category (spectral peak in the theta [4-8 Hz] range) corresponds to a severely disconnected system wherein cortical neurons are comparatively more depolarized resulting in theta frequency bursting.^9^ The ‘C’ category (spectral peaks in the theta and beta [13-24 Hz] ranges) corresponds to a moderately disconnected system wherein thalamic neuron bursting produces coexisting theta and beta frequency oscillations in connected cortex.^10,11^ Finally, the ‘D’ category (spectral peaks in the alpha [8-13 Hz] and beta ranges) corresponds to a fully intact thalamocortical network wherein a sufficiently structurally and functionally interconnected thalamus and cortex allows for production of alpha and beta frequency oscillations.^12^

Because the ABCD framework provides a means for inferring thalamocortical functional integrity from EEG dynamics, it has potential to aid clinicians in the assessment of acute severe brain injury. The intensive care unit (ICU) setting poses a number of challenges to patient assessment including sedation, medical comorbidities, a high rate of misdiagnosis with behavioural exams, and the potential presence of cognitive motor dissociation (CMD).^13-15^ Therefore, resting-state measures that can be gleaned from recordings collected during routine clinical care with widely available equipment, such as EEG, may serve as useful complements to advanced task- and stimulus-based assessments (e.g., EEG and functional MRI [fMRI] command-following and passive language paradigms)^16-29^ as well as allow for higher sampling frequencies and dynamic tracking in a population that experiences unpredictable arousal fluctuations.^17,30-35^

However, prior to its implementation in the clinical setting, the generalizability of and optimal method of applying the ABCD framework need to be understood. To date, this approach has only been studied in post-cardiac arrest coma^7^ and a heterogeneous sample of patients with acute non-hypoxic-ischemic brain injury.^36^ Moreover, its utility in analyzing EEG topography has not been explored in prior studies wherein each participant was assigned to a single ABCD category based on visual inspection of either all EEG channels simultaneously^7^ or a single channel.^36^ While these approaches may be suitable for patients with uniform injury burden across the cerebrum (such as in hypoxic-ischemic injury), patients with traumatic brain injury (TBI) present with heterogeneous, multifocal disruptions of the thalamocortical network caused by axonal shearing injury.^37^ As a result, portions of the thalamocortical network are often preserved, even in patients with severe injuries who present in coma.^38^

In the present study, we applied the ABCD framework to investigate channel-level EEG signal dynamics in a sample of patients with acute severe TBI to determine the topographic distribution of ABCD classifications across patients. We studied the within-session temporal stability of ABCD classifications during a single, 2-hour data collection session. In a subset of patients, we investigated the longitudinal evolution of EEG signals at ≥6-months post-injury to elucidate how ABCD classifications change with recovery. We tested four hypotheses: 1) given the well-established spatial heterogeneity of thalamocortical disconnection and susceptibility to state fluctuations in severe TBI, patients exhibit spatial heterogeneity and within-session temporal variability in channel-level ABCD classifications; 2) ABCD classifications improve longitudinally after severe TBI, commensurate with the degree of behavioural recovery; 3) ABCD classifications correlate with behavioural level of consciousness in the acute and subacute-to-chronic phases of recovery; and 4) administration of the Coma Recovery Scale-Revised (CRS-R) arousal facilitation protocol, which is a commonly used clinical maneuver used to promote arousal in DoCs,^39^ yields improved EEG dynamics along the ABCD scale. Our aim was to investigate the characteristics of EEG signals in the context of the ABCD framework so as to inform future clinical implementation of this approach for patients with severe TBI.

## Materials and methods

### Participants

We enrolled 20 patients who presented with acute traumatic coma to the ICU at an academic medical center between March 2012 and January 2017 (median age: 26.5 years, interquartile range [IQR]: 7.8 years, 14 males), 12 of whom were studied longitudinally at ≥6-months post-injury (median: 215 days; range: 160-1172 days) (Table 1). We attempted to contact all surviving patients or their surrogates for follow-up. Our study cohort included a previously published cohort of 16 patients prospectively enrolled in the ICU (P1-16),^23^ as well as a convenience sample of four additional patients that were enrolled at ≥6-month follow-up (P17-20) after being referred to an investigator’s NeuroRecovery Clinic (B.L.E.). For the four enrolled at follow-up,^40^ their acute clinical EEG and behavioural data were obtained retrospectively.

**Table 1.** Patient demographics and clinical information.

| Patient ID | Age range at injury (years) | Sex | TBI mechanism | iGCS | Acute/ Follow-Up | Day of EEG | LoC at EEG | Total CRS-R at EEG | CRS-R subscale scores at EEG |
| --- | --- | --- | --- | --- | --- | --- | --- | --- | --- |
| P1 | 23-27 | M | MVA | 5T | Acute<br>Follow-Up | 17<br>206 | PTCS<br>R-PTCS | 23<br>23 | A4V5M6O3C2Ar3<br>A4V5M6O3C2Ar3 |
| P2 | 18-22 | M | Ped versus car | 4-8T | Acute<br>Follow-Up | 2<br>174 | MCS-<br>R-PTCS | 4<br>23 | A0V0M3O1C0Ar0<br>A4V5M6O3C2Ar3 |
| P3 | 18-22 | F | MVA | 5T | Acute<br>Follow-Up | 4<br>371 | Coma<br>R-PTCS | 1<br>23 | A0V0M1O0C0Ar0<br>A4V5M6O3C2Ar3 |
| P4 | 18-22 | M | Fall | 3-7T | Acute<br>Follow-Up | 10<br>576 | PTCS<br>R-PTCS | 22<br>23 | A4V5M6O3C2Ar2<br>A4V5M6O3C2Ar3 |
| P5 | 33-37 | M | Fall | 5T | Acute | 16 | VS | 3 | A0V0M0O2C0Ar1 |
| P6 | 28-32 | F | MVA | 3 | Acute<br>Follow-Up | 10<br>656 | MCS+<br>R-PTCS | 11<br>23 | A3V2M3O1C0Ar2<br>A4V5M6O3C2Ar3 |
| P7 | 43-47 | M | MVA | 5T | Acute | 14 | MCS+ | 18 | A3V5M5O3C1Ar1 |
| P8 | 33-37 | M | Fall | 5-7T | Acute | 8 | PTCS | 20 | A4V5M5O2C2Ar2 |
| P9 | 28-32 | M | Ped versus car | 5-7T | Acute | 13 | MCS+ | 15 | A3V3M5O1C1Ar2 |
| P10 | 23-27 | M | Assault | 3-7T | Acute | 13 | MCS- | 10 | A1V1M5O1C0Ar2 |
| P11 | 18-22 | F | Ped versus car | 6T | Acute<br>Follow-Up | 14<br>187 | PTCS<br>R-PTCS | 22<br>23 | A4V5M6O3C1Ar3<br>A4V5M6O3C2Ar3 |
| P12 | 23-27 | F | Fall | 3 | Acute | 8 | Coma | 1 | A0V0M1O0C0Ar0 |
| P13 | 18-22 | M | Fall | 3-7 | Acute | 6 | PTCS | 21 | A4V5M6O3C2Ar1 |
| P14 | 28-32 | M | Ped versus car | 4-7 | Acute<br>Follow-Up | 8<br>235 | MCS+<br>R-PTCS | 7<br>23 | A3V0M3O0C0Ar1<br>A4V5M6O3C2Ar3 |
| P15 | 33-37 | M | Fall | 3-4 | Acute<br>Follow-Up | 4<br>191 | PTCS<br>R-PTCS | 14<br>23 | A4V2M6O0C0Ar2<br>A4V5M6O3C2Ar3 |
| P16 | 23-27 | M | Fall | 4 | Acute | 28 | PTCS | 18 | A3V4M6O2C1Ar2 |
| P17 | 23-27 | F | Ped versus car | 3-5T | Acute<br>Follow-Up | 4<br>224 | Coma<sup>a</sup><br>MCS- | N/A<br>9 | N/A<br>A1V3M2O1C0Ar2 |
| P18 | 23-27 | M | MVA | 3-6T | Acute<br>Follow-Up | 5<br>182 | Coma<sup>a</sup><br>MCS+ | N/A<br>15 | N/A<br>A4V3M3O2C1Ar2 |
| P19 | 18-22 | F | Ped versus car | 3 | Acute<br>Follow-Up | 4<br>160 | Coma<sup>a</sup><br>VS | N/A<br>5 | N/A<br>A2V0M1O1C0Ar1 |
| P20 | 28-32 | M | MVA | 3-5T | Acute<br>Follow-Up | 14<br>1172 | Coma<sup>a</sup><br>MCS- | N/A<br>5 | N/A<br>A0V0M3O1C0Ar1 |

Initial Glasgow Coma Scale (iGCS) is defined as the highest and lowest post-resuscitation GCS score assessed by a qualified clinician and not confounded by sedation or paralytics prior to ICU admission. Level of consciousness (LoC) was assessed with either the Coma Recovery Scale – Revised (CRS-R) (P1-16, all patients at follow-up) or neurological examination (P17-20 acutely) as coma, vegetative state (VS), minimally conscious state with/without language (MCS+/-), post-traumatic confusional state (PTCS), or recovered from post-traumatic confusional state (R-PTCS). CRS-R subscales are as follows: Auditory Function (A), Visual Function (V), Motor Function (M), Oromotor Function (O), Communication (C), and Arousal (Ar).
F = female; M = male; MVA = motor vehicle accident; N/A = not applicable; Ped = pedestrian; TBI = traumatic brain injury.
<sup>a</sup>LoC derived from clinical neurological examination rather than CRS-R.

Inclusion criteria were: 1) age 18-65 years; and 2) head trauma with Glasgow Coma Scale (GCS) score of 3-8 and no eye opening for ≥24 hours. Exclusion criteria were: 1) life expectancy <6 months, as estimated by a treating physician; 2) prior neurodegenerative disease or severe brain injury; 3) body metal precluding MRI; and 4) no fluency in English prior to injury (because the task- and stimulus-based paradigms administered as part of the study protocol were administered in English).^23^ Given that this study was a retrospective analysis of all data collected in a previously published study^23^ and that previously published data on the ABCD classification framework are insufficient to inform a power calculation, we included all patients with resting-state EEG data available to us. We also enrolled 16 healthy control participants (median age: 27 years, IQR: 21-32.5 years, 12 males). All studies were approved by the Mass General Brigham Institutional Review Board. We obtained written informed consent from healthy control participants and from surrogate decision-makers of patients with DoCs. Patients who recovered consciousness at follow-up provided informed consent for their continued participation in the study.

### Neurobehavioural assessments

To evaluate behavioural level of consciousness, we either administered the CRS-R (P1-16 acutely and all patients at follow-up)^39^ or derived level of consciousness from a clinical bedside neurological exam conducted on the same day as each EEG (P17-20 acutely). We graded the level of consciousness as coma, vegetative state (VS), minimally conscious state with/without language function (MCS+/-) or emerged from the minimally conscious state (EMCS). We used the Confusion Assessment Protocol (CAP)^41^ to confirm whether patients who emerged from MCS were in a post-traumatic confessional state (PTCS) or recovered from PTCS (R-PTCS).^42^ A single investigator (B.L.E.) conducted all CRS-R and CAP assessments, blinded to the EEG data. Acute clinical neurological exams were conducted by treating physicians.

### EEG data acquisition and preprocessing

We acquired two to five 5-minute blocks of resting-state EEG from all participants. Sedative, anxiolytic, and/or analgesic medications were administered to a subset of patients before and/or during the acute EEG recording at the discretion of the clinical team for patient safety or comfort (Supplementary Table 1). For healthy control participants, patients P1-16 acutely, and all patients studied at follow-up, rest blocks were interspersed within a previously published 2-hour battery of task- and stimulus-based EEG paradigms (e.g., motor imagery, language, and music).^23^ The CRS-R arousal facilitation protocol, which is designed to promote wakefulness via sustained deep pressure stimulation of face, sternocleidomastoid, trapezius, arm, and leg muscles,^39^ was administered immediately prior to the penultimate rest block in a subset of patients studied acutely (*n* = 12) and at follow-up (*n* = 10) that were able to complete the full study protocol. During EEG collection, participants in this subset were instructed to keep their eyes closed (to maintain consistency with participants that could not open their eyes) and were not disturbed by members of the investigational team, with the exception of administration of the CRS-R arousal facilitation protocol between rest blocks four and five. We did not screen for sleep because we aimed to maintain consistency across participants given the lack of reliable electrophysiologic indicators of sleep in acute brain injury.^43^

For patients P17-20, we obtained EEG from the acute hospitalization by selecting the earliest 5-minute clinical EEG segments with minimal ambient noise and artefact that immediately followed auditory or tactile stimulation (e.g., from routine nursing care) to increase the likelihood of capturing data from periods of maximal patient level of arousal.^28^

We collected EEG with a Natus XLTEK EEG system (San Carlos, CA) with 19 electrodes arranged via a 10-20 system (200, 250 or 256 Hz sampling frequency).^44^ Data were detrended, filtered (third-order Butterworth, zero-phase shift digital filter [1-30 Hz]), cut into 3-second non-overlapping epochs, and re-referenced to the Hjorth Laplacian montage to increase our ability to localize the sources of recorded signals.^45,46^ We performed all data processing in MATLAB (The Mathworks, Natick, MA) using a combination of EEGLAB and the Chronux toolbox.^47,48^

We visually inspected signals and rejected epochs and, if necessary, entire rest blocks, contaminated by electromyogenic, eye-blink, or electrical interference artefact. A single investigator (W.H.C.) blinded to clinical and behavioural assessment data performed all manual artefact rejection to ensure consistency across blocks.^7,17,49^ Manual artefact rejection resulted in an average of 40.2% of 3-second epochs rejected for healthy controls and 60.9% of 3-second epochs rejected for patients, consistent with previously reported artefact rejection rates for this method.^17^

### EEG spectral analysis and ABCD classification

We calculated channel-level power spectral densities for each 3-second epoch with the multitaper method using the Chronux toolbox (five tapers, yielding a frequency resolution of 2 Hz and estimates spaced 1/3 Hz apart) and then averaged these spectra within each rest block.^48,50,51^ To assess the effect of the CRS-R arousal facilitation protocol on ABCD classifications for the subset of patients that it was performed on (*n* = 12 patients acutely, *n* = 10 patients at follow-up), we computed spectra from the two minutes immediately following the completion of the arousal facilitation protocol (given the transient nature of the protocol’s effect on arousal) and compared the corresponding ABCD classifications (assigned as described below) to the ABCD classifications from the spectra calculated from the 5-minute rest block immediately preceding the arousal facilitation protocol (block four).

For each rest block, we assigned each channel’s EEG recording to a pre-defined ABCD category via visual inspection of its spectrum.^2,7,36^ The criteria were as follows: an ‘A’-type spectrum either lacked any spectral peaks or contained a delta (<4 Hz) frequency peak; a ‘B’- type spectrum contained only a theta (4-8 Hz) frequency peak; a ‘C’-type spectrum contained both theta and beta (13-24 Hz) frequency peaks; and a ‘D’-type spectrum contained both alpha (8-13 Hz) and beta frequency peaks. For spectra that met multiple requirements, the most favorable category (D>C>B>A) was assigned. We rejected spectra with evidence of electromyogenic artefact contamination and left those recordings unscored. Spectra that did not fit into any category (e.g., presence of an alpha peak only) were also unscored. To quantify ABCD classifications for each participant, we coded channel-level ABCD classifications (omitting the unscored recordings) as numeric values (A = 1, B = 2, C = 3, D = 4) and then derived an ‘ABCD index’ by averaging across channels.

To mitigate potential bias in the visual assessment of spectral peaks that might be associated with knowledge of the participant, cohort (healthy control, acute patient, follow-up patient) or channel location, we performed ABCD classification on spectra that were randomly shuffled without any identification of participant, cohort, channel, or block. As previously reported by Forgacs and colleagues,^7^ this method of visual inspection-based classification has a high inter-rater reliability (89% concordance). Thus, ABCD spectral classification was conducted by a single investigator (W.H.C.) who did not have patient contact, was not involved in data collection, and was blinded to all participant identifiers, channel information, and clinical variables. This investigator was also one of the raters in the previous study demonstrating high inter-rater reliability of visual inspection-based classification of ABCD spectra.^7^

### Statistical analyses

All statistical analyses were conducted with Prism 9.0 (GraphPad Software, San Diego, CA) and *P*-values < 0.05 were considered statistically significant. To test whether the ABCD index and percentages of individual ABCD classifications changed between acute ICU hospitalization and follow-up, we used Wilcoxon matched-pairs signed rank tests (two-tailed). We calculated Mann-Whitney statistics (two-tailed) to investigate the difference in ABCD indices from patients with and without command-following.

To assess the within-session temporal stability of ABCD classifications, we calculated block-level ABCD indices for each participant. For each participant with five rest blocks recorded during a continuous 2-hour window (*n* = 13 healthy controls, *n* = 10 acute and follow-up patients), we calculated the root mean square deviation (RMSD) of the ABCD index across rest blocks as a measure of inter-block variability. We used a Wilcoxon matched-pairs signed rank test (two-tailed) to assess the difference in acute versus follow-up patient RMSDs and Mann-Whitney statistics (two-tailed) to assess the difference in control versus acute patient and control versus follow-up patient RMSDs.

To test the hypothesis that the CRS-R arousal facilitation protocol increases the ABCD index in acute (*n* = 12) and follow-up (*n* = 10) patients, we used Wilcoxon matched-pairs signed rank tests (one-tailed) to compare the ABCD index from the rest block preceding the CRS-R arousal facilitation protocol to the ABCD index from the 2 minutes immediately following the completion of the arousal facilitation protocol.

### Data availability

The data that support the findings of this study are available upon reasonable request from the corresponding author and approval from the Institutional Review Board.

## Results

### Patient demographics

The patient cohort studied acutely consisted of 20 individuals with the following behavioural diagnoses at time of acute EEG collection: coma (*n* = 6), VS (*n* = 1), MCS- (*n* = 2), MCS+ (*n* = 4), PTCS (*n* = 7) (Table 1). All patients assessed with clinical neurological exam rather than CRS-R (P17-20) were in coma at the time of acute EEG. Median time from injury to acute EEG was 9 days (range: 2-28 days). Patients P1-16 were enrolled consecutively as part of a previously published study (399 consecutive patients screened for eligibility, 28 patients met all eligibility criteria, 16 patients were enrolled),^23^ while patients P17-20 represent a convenience sample enrolled at follow-up. Sedative, analgesic, and anxiolytic medications administered before and/or during EEG are reported in Supplementary Table 1. In total, six patients received continuous infusions of sedatives and 11 patients received intravenous boluses of enteral analgesics or anxiolytics before and/or during the acute EEG. Twelve patients were studied at ≥6-month follow-up (median: 215 days; range: 160-1172 days) and had the following behavioural diagnoses: VS (*n* = 1), MCS- (*n* = 2), MCS+ (*n* = 1), R-PTCS (*n* = 8). Eight patients did not complete follow-up for one of the following reasons: they were deceased, they were unable to participate because of logistical and/or ongoing medical issues, or they (or their surrogate) declined to participate further.

### ABCD-classified EEG signal characteristics

Overall, EEG data from all participants were successfully classified into ABCD categories. We classified 86.9% and 75.3% of channel-level spectra from controls and patients into ABCD categories, respectively. Spectra that were not classified either contained spectral peak patterns not consistent with any of the ABCD categories (11.9% for controls, 20.2% for patients) or were rejected because of artefacts (1.2% for controls, 4.5% for patients). Of classifiable channels, healthy participants demonstrated ‘D’ signals in an average of 93.7% of channels, and 75.0% (12/16) of healthy participants demonstrated ‘D’ signals in all channels. The majority (95.0%) of patients demonstrated ‘D’ signals in one or more channels acutely, but there was marked inter-patient heterogeneity in both the proportion and location of different ABCD classifications (Fig. 1). Patients studied acutely demonstrated ‘D’ signals in a mean 37.4% of classified channels (range: 0 – 93.9%, standard deviation (SD): 31.6%), ‘C’ signals in 28.0% (range: 0 – 90.5%, SD: 23.0%), ‘B’ signals in 22.6% (range: 0 – 80.0%, SD: 26.6%), and ‘A’ signals in 12.0% (range: 0 – 57.5%, SD: 15.9%). Visual inspection of channel-level ABCD classifications revealed homogeneity throughout the brain in a subset of patients (e.g., P2), while others demonstrated coexistence of multiple disparate ABCD classifications (e.g., P20). By contrast, healthy controls generally had ‘D’ signals in most or all channels (Fig. 1).

**Figure 1.**
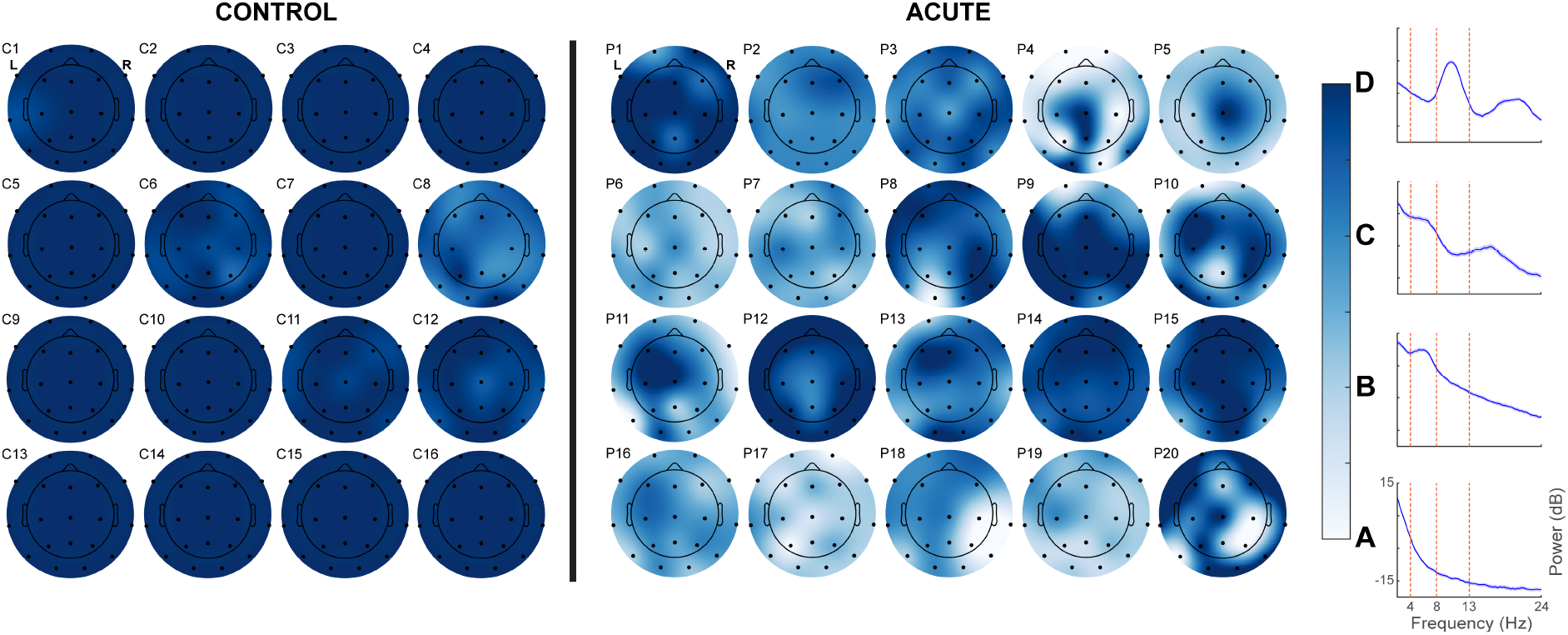
Heterogeneity of ABCD-classified EEG signals in patients with acute severe TBI. Channel-level averages of ABCD classifications across all rest blocks for all healthy controls (*n* = 16) and patients studied acutely (*n* = 20). Heatmaps indicate classification (D = 4, A = 1), according to electrode location on the scalp (represented by black dots). Missing electrodes for some participants represent channels that were omitted on the basis of artefact or inability to be classified into an ABCD category. Representative spectra for each of the ABCD categories are shown to the right of the colour bar. dB = decibels, Hz = Hertz.

### Within-session temporal stability of ABCD classifications

We utilized the ABCD framework to assess within-session fluctuations of EEG dynamics (Fig. 2) by examining how classifications changed within a recording session among participants who had five discrete rest blocks recorded within a 2-hour window (*n* = 13 healthy controls, *n* = 10 acute and follow-up patients). In the acute recordings, there was significantly higher inter-block variability relative to healthy controls (difference of medians: 0.3, 95% confidence interval [CI]: 0.07-0.4, *P* = 0.005). There was no significant difference in inter-block variability in acute recordings relative to follow-up recordings (difference of medians: 0.1, 95% CI: -0.4-0.2, *P* = 0.2) or in follow-up recordings relative to healthy control recordings (difference of medians: 0.09, 95% CI: 0-0.2, *P* = 0.5). Thirty-five percent (7/20) of acute patients demonstrated an inter-block ABCD index range ≥ 1 (corresponding to the interval between ABCD categories), while the same was true for only 6.3% (1/16) of controls and 8.3% (1/12) of follow-up patients (Fig. 2B-C). Additionally, all controls demonstrated ‘D’-type signals in all channels during the first rest block (block one) and the last rest block (block five), which immediately followed administration of the CRS-R arousal facilitation protocol (Fig. 2B).

**Figure 2.**
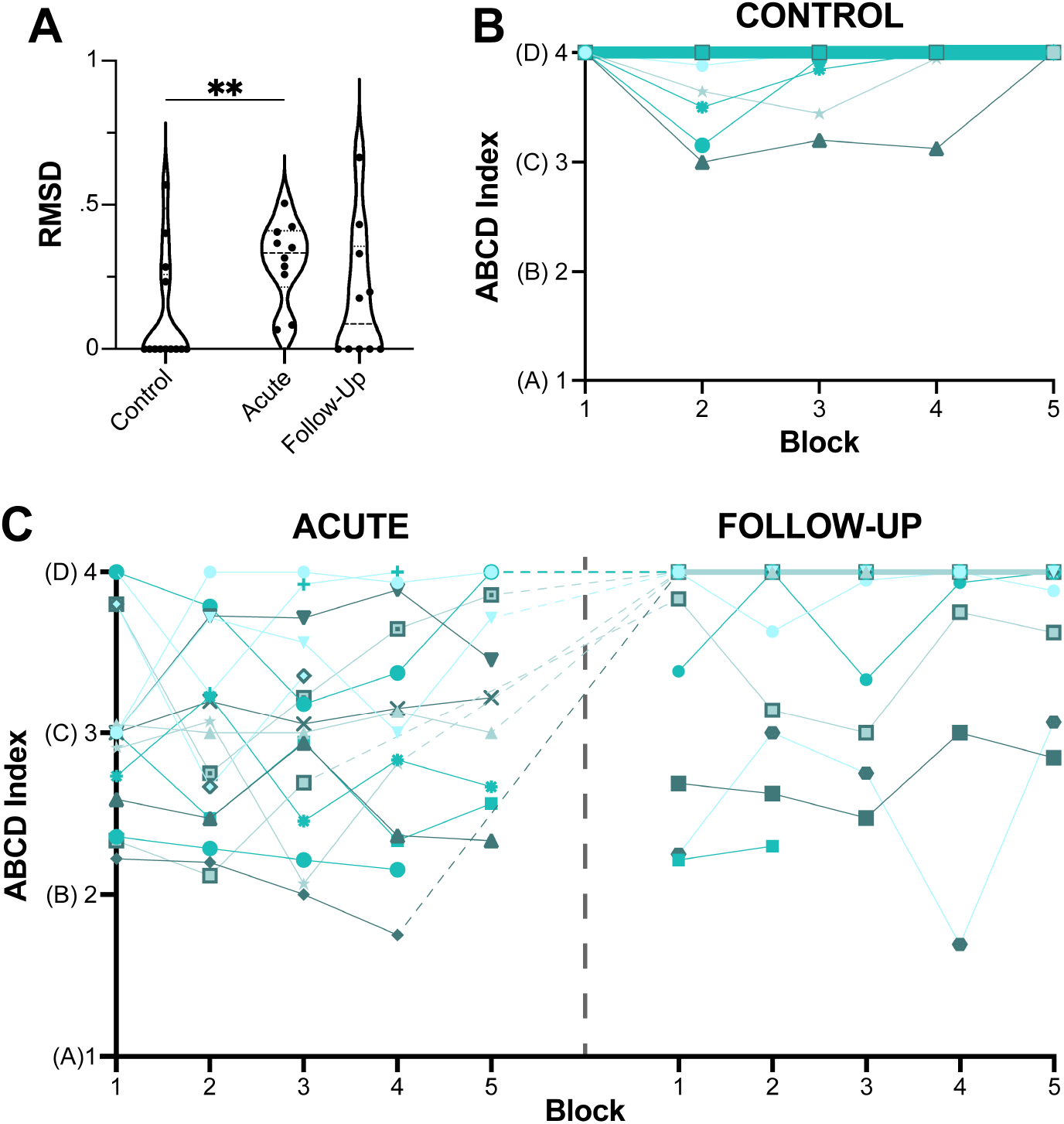
Within-session temporal stability of ABCD classifications. (**A**) Root mean square deviation (RMSD) of ABCD indices calculated across rest blocks for the subset of participants who had five discrete rest blocks collected within a contiguous 2-hour window (*n* = 16 healthy controls, *n* = 10 patients). Patients studied acutely demonstrated significantly higher RMSDs relative to healthy controls (difference of medians: 0.3, *P* = 0.005). Within violin plots, dashed lines indicate the median and dotted lines indicate the 25^th^ and 75^th^ percentiles. (**B**) ABCD indices for each rest block for all healthy controls (*n* = 16). (**C**) ABCD indices for each rest block for patients studied acutely (*n* = 16) and at ≥6-month follow-up (*n* = 12) with discrete rest blocks recorded at regularly spaced intervals. For **B** and **C**, each participant is represented by a line/symbol and, for overlapping lines, line thickness is proportional to the number of participants. \*\**P*<0.01 (Mann-Whitney test).

### Longitudinal changes in ABCD dynamics

In Fig. 3A, we show the ABCD index calculated across all channels and rest blocks for each patient studied acutely and at follow-up (an interval of at least 6 months). ABCD indices significantly increased between the acute ICU hospitalization and follow-up among the 12 patients studied longitudinally (median change: 0.6, 95% CI: 0.3-1.0, *P* = 0.006). We also assessed the change in the percentage of different ABCD categories for each patient (Fig. 3B) and found that, on a group level, the proportion of ‘D’ signals increased between the acute ICU hospitalization and follow-up (median change: 36.7%, 95% CI: 4.2-72.5%, *P* = 0.02), while the proportion of ‘A’ signals decreased (median change: -5.3%, 95% CI: -16.9-0%, *P* = 0.008). As the increase in the fraction of ‘D’ signals was greater than the decrease in the fraction of ‘A’ signals, the proportion of ‘B’ and ‘C’ signals also decreased, but these differences were not statistically significant. Figure 3C shows subject-level topographic plots averaged across all rest blocks in the acute ICU setting and at follow-up with patients stratified by behavioural diagnosis at follow-up, corroborating the increase in ‘D’ classifications at follow-up versus the acute recording shown in Figure 3B. Figure 3C also shows that ‘D’ classifications were more common in patients with higher behavioural levels of consciousness, which we investigate below.

**Figure 3.**
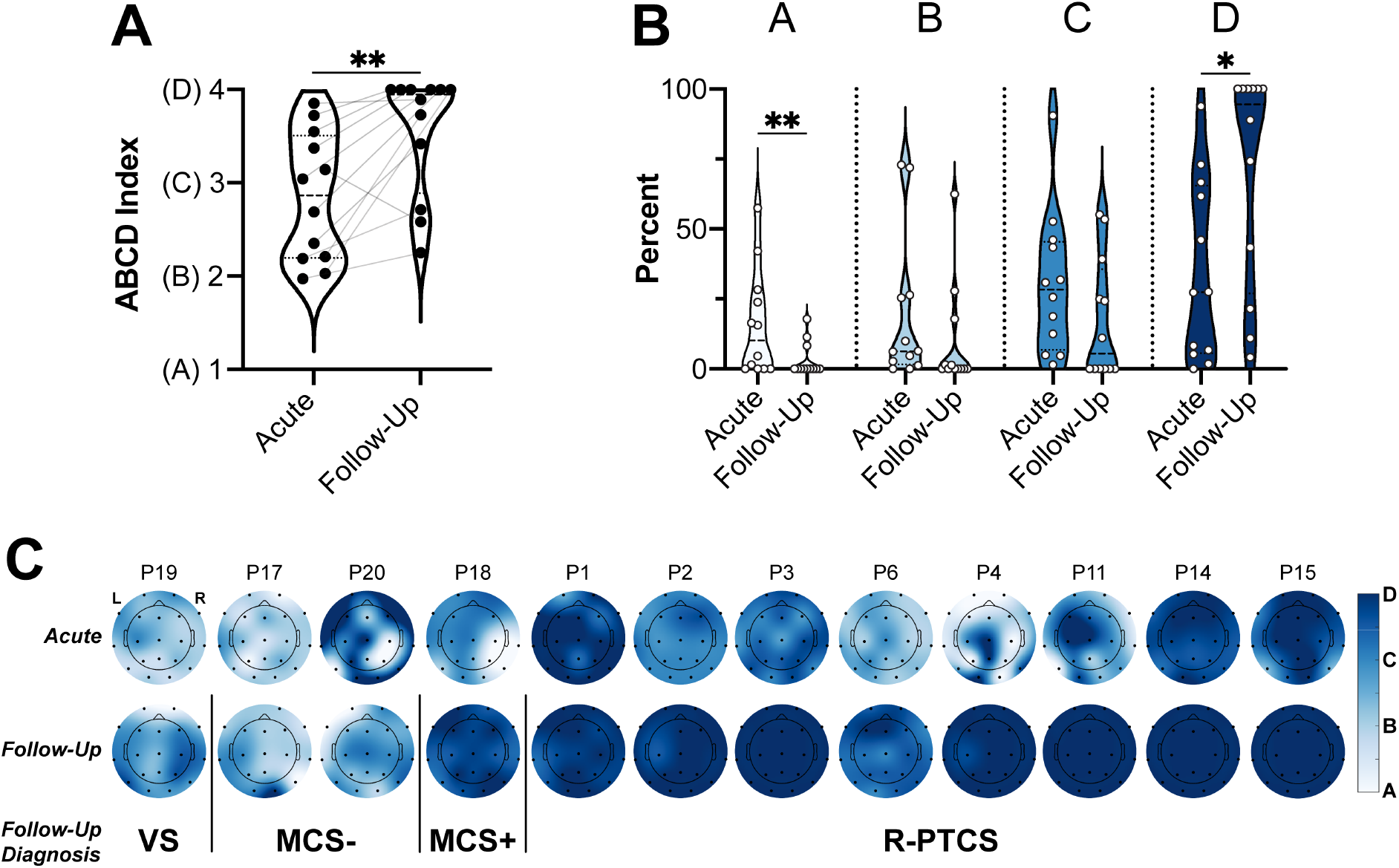
Longitudinal changes in ABCD dynamics. (**A**) ABCD index averaged across all channels and rest blocks for each patient studied both acutely in the ICU and at ≥6-month follow-up (*n* = 12). Patients demonstrated significantly higher ABCD indices at follow-up as compared to the acute setting (median change: 0.6, *P* = 0.006). Lines connect sequential recordings from individual patients. (**B**) Percent of channels classified into each of the ABCD categories (of the channels that could be classified) for each patient studied both acutely and at follow-up (*n* = 12). Follow-up patients demonstrated significantly increased proportions of ‘D’-type signals (median change: 36.7%, *P* = 0.02) and significantly decreased proportions of ‘A’- type signals (median change: -5.3%, *P* = 0.008). (**C**) Channel-level averages of ABCD classifications across all rest blocks for patients studied both acutely and at follow-up (*n* = 12). Patients are ordered according to behavioural diagnosis at ≥6-month follow-up. Heatmaps indicate classification (D = 4, A = 1), according to electrode location on the scalp (represented by black dots). Missing electrodes for some participants represent channels that were omitted on the basis of artefact or inability to be classified into an ABCD category. For **A** and **B**, dashed lines within violin plots indicate the median and dotted lines indicate the 25^th^ and 75^th^ percentiles. \**P*<0.05, \*\**P*<0.01 (Wilcoxon matched-pairs signed rank test).

### Association of ABCD classifications with command-following and behavioural diagnosis

Eleven of 20 acute patients and nine of 12 follow-up patients demonstrated behavioural command-following at the time of EEG (Fig. 4A). Given the presence of rapid state fluctuations among patients studied acutely (Fig. 2), we assessed whether patients’ ‘best ABCD index,’ defined as the highest ABCD index across all recorded rest blocks, was associated with behavioural level of consciousness at the same timepoint (Fig. 4A). There was no difference in the best ABCD index for acute patients with behavioural command-following as compared to those without (difference of medians: 0.09, 95% CI: -0.9-0.4, *P* = 0.4). By contrast, follow-up patients with behavioural command-following demonstrated significantly higher best ABCD indices as compared to those without (difference of medians: 1.0, 95% CI: 0.9-1.7, *P* = 0.005). Of note, we observed that all follow-up patients diagnosed as MCS+ or R-PTCS (*n* = 9) demonstrated best ABCD indices ≥ 3.8 while all follow-up patients diagnosed as VS or MCS- (*n* = 3) demonstrated best ABCD indices ≤ 3.1 (Fig. 4B).

**Figure 4.**
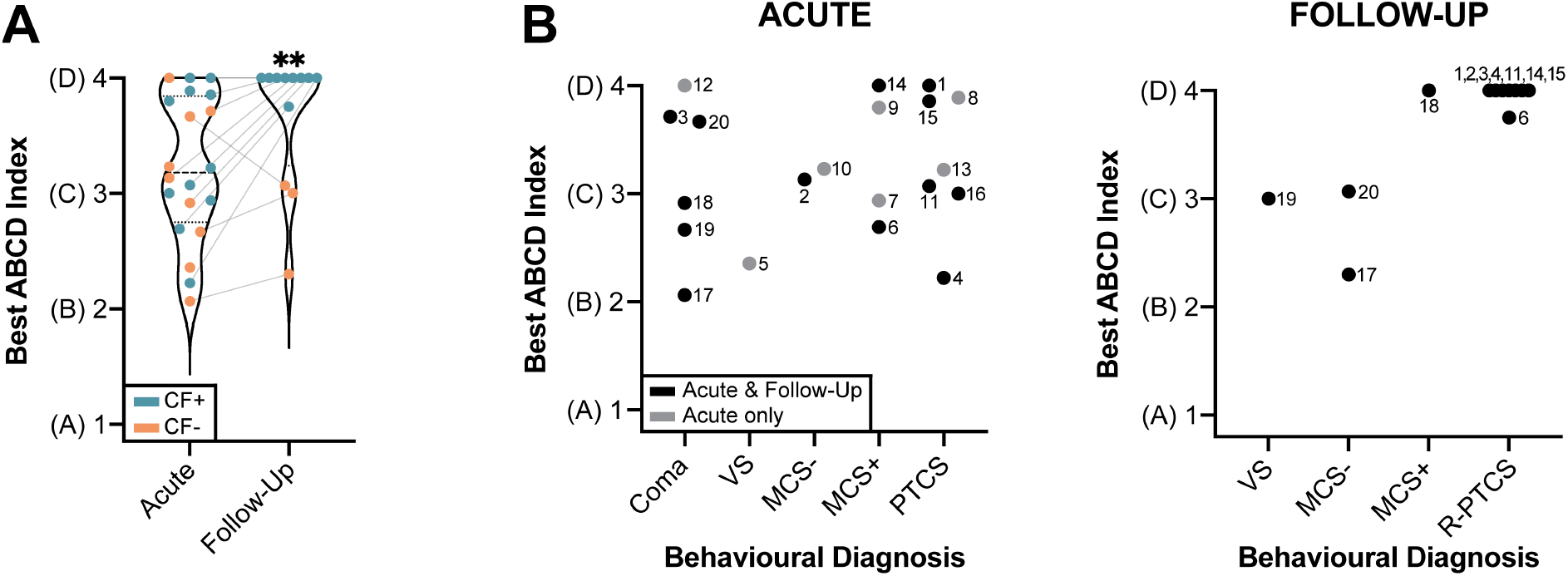
Comparison of ABCD classification and behavioural diagnosis. (**A**) Best ABCD index (defined as the highest ABCD index across all rest blocks) for all acute (*n* = 20) and follow-up (*n* = 12) patients with (blue; CF+) and without (orange; CF-) behavioural evidence of command-following. Lines connect sequential recordings from patients studied both acutely and at follow-up (*n* = 12). There was no difference in the best ABCD index for the acute patients with command-following (*n* = 11) as compared to those without (*n* = 9) (difference of medians: 0.09, 95% CI: -0.9-0.4, *P* = 0.4), while follow-up patients with command-following (*n* = 9) demonstrated significantly higher best ABCD indices as compared to follow-up patients without (*n* = 3) (difference of medians: 1.0, 95% CI: 0.9-1.7, *P* = 0.005). Within violin plots, dashed lines indicate the median and dotted lines indicate the 25^th^ and 75^th^ percentiles. (**B**) Best ABCD index for all patients plotted according to behavioural diagnosis. Patients studied longitudinally are plotted in black and patients studied in the acute ICU phase only are plotted in gray. Numbers adjacent to dots represent patient identifiers. \*\**P*<0.01 (Mann-Whitney test).

In acute recordings, we observed a range of best ABCD indices for patients with behavioural diagnoses of coma, MCS+, and PTCS (Fig. 4B). Two of the three longitudinally studied patients with acute best ABCD indices > 3 but without behavioural evidence of command-following ultimately recovered to R-PTCS at follow-up (P2, P3) while the remaining patient was diagnosed as MCS-at follow-up (P20) and demonstrated a decrease in the best ABCD index over time. One of the three longitudinally studied patients with an acute best ABCD index < 3 recovered to MCS+ at follow-up (P18) while the others were diagnosed as VS (P19) and MCS- (P17) at follow-up.

### Effect of CRS-R arousal facilitation protocol on ABCD classification

To test the hypothesis that the CRS-R arousal facilitation protocol yields an improvement in EEG signal dynamics, we compared the ABCD index in the 5-minute rest block preceding the arousal facilitation protocol to the ABCD index in the 2 minutes immediately following (Fig. 5). Patients studied acutely demonstrated significantly higher ABCD indices following the CRS-R arousal facilitation protocol (*n* = 12, median of differences: 0.1, 95% CI: -0.05-0.6, *P* = 0.03), while follow-up patients demonstrated no change (*n* = 10, median of differences: 0, 95% CI: -0.08-0, *P* = 0.4). Of note, the lack of a change in the follow-up recordings may be due to a ceiling effect, as six of 10 patients had ABCD indices > 3.9 both prior to and after the CRS-R arousal facilitation protocol.

**Figure 5.**
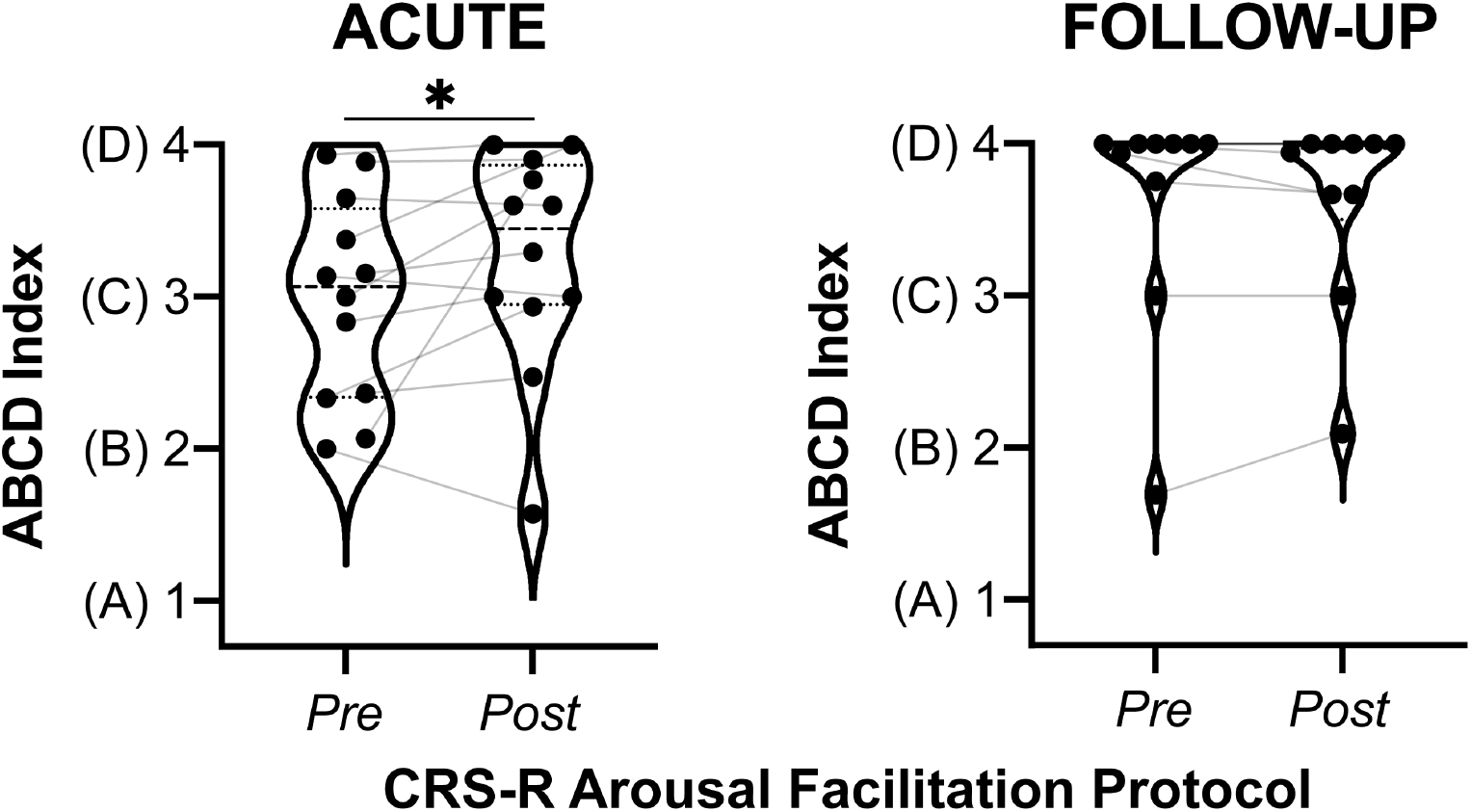
Effect of CRS-R arousal facilitation protocol on ABCD classification. ABCD index calculated from the rest block immediately preceding (*Pre*) and the 2 minutes immediately following (*Post*) the CRS-R arousal facilitation protocol in acute (*n* = 12) and follow-up (*n* = 10) recordings. The CRS-R arousal facilitation protocol involves sustained, rhythmic application of deep pressure stimulation to the face, sternocleidomastoid, trapezius, arm, and leg muscles, and is designed to promote arousal in patients with DoCs. In acute recordings, ABCD indices were significantly higher following administration of the CRS-R arousal facilitation protocol (median of differences: 0.1, 95% CI: -0.05-0.6, *P* = 0.03). In follow-up recordings, ABCD indices did not significantly increase following administration of the CRS-R arousal facilitation protocol (median of differences: 0, 95% CI: -0.08-0, *P* = 0.4). Six of 10 follow-up patients had ABCD indices > 3.9 both prior to and after the CRS-R arousal facilitation protocol. Within violin plots, dashed lines indicate the median and dotted lines indicate the 25^th^ and 75^th^ percentiles. \**P*<0.05 (Wilcoxon matched-pairs signed rank test).

## Discussion

In this longitudinal study of patients with acute severe TBI, we analyzed EEG dynamics at the level of individual channels. Our analysis is based on the mesocircuit model for DoCs,^1^ which proposes that different levels of functional thalamocortical network integrity correspond to specific EEG dynamics.^2,7^ Because these distinct dynamical patterns are manifest by characteristic peaks in the power spectrum, EEG signals can be visually classified as ‘A,’ ‘B,’ ‘C,’ or ‘D,’ corresponding to the range of thalamocortical network functioning from absent to normal. We found that while all patients with acute severe TBI, as expected, had EEG signal characteristics indicating dysfunction of the thalamocortical network, the majority also demonstrated channel-level EEG signals consistent with at least partial thalamocortical network preservation. Moreover, there was marked heterogeneity in both the proportion and location of ABCD classifications across patients. Acutely injured patients demonstrated significantly more within-session temporal variability in EEG signals in comparison to healthy control participants. Furthermore, over a third of acute patients demonstrated a within-session ABCD index fluctuation ≥ 1 (the interval between ABCD categories) within a single 2-hour window, highlighting the need to perform multiple assessments of thalamocortical network function in patients with acute severe brain injuries. As a group, patients studied at ≥6-month follow-up demonstrated significant improvement along the ABCD scale in comparison to the acute phase of injury, driven by an increase in ‘D’-type signals and a decrease in ‘A’-type signals. At follow-up, patients with the ability to follow commands also demonstrated higher ABCD indices as compared to those without. Importantly, we found that the CRS-R arousal facilitation protocol yielded improved EEG signal dynamics in the context of the ABCD framework, providing electrophysiological evidence for the utility of this widely used behavioural examination technique. Collectively, these findings support the use of the ABCD framework to characterize channel-level EEG dynamics and thereby track fluctuations in thalamocortical network function in the ICU and other settings.

### State fluctuations in acute severe TBI

We found that in acute recordings, there was significantly more within-session temporal variability in ABCD states relative to control recordings (Fig. 2). Furthermore, over a third of acute patients demonstrated a within-session change in the ABCD index ≥ 1, indicating a global shift from one state of thalamocortical functioning to another within a single 2-hour window. This observation bolsters the notion that patients with severe brain injuries exhibit marked fluctuations in state within short time periods and therefore require repeated assessment to yield accurate diagnoses.^15,32-35,52-54^

Given the ease of deriving ABCD classifications from clinical EEG without requiring investigators at the bedside, ABCD classification may provide clinicians with the ability to sample patients’ thalamocortical network functioning at a frequency not attainable with other tools. Furthermore, state fluctuations may influence a patient’s ability to demonstrate a positive response on task-based assessments.^17^ Thus, a real-time application of ABCD classification may allow for identification of periods of elevated arousal to guide the timing of task-based measures so as to maximize the chance of detecting positive responses in covertly conscious patients. With replication, it may also be used as a validity indicator for standardized behavioural assessments, such as the CRS-R.^39^

ABCD-based timing of behavioural assessment and task-based fMRI and EEG may be particularly relevant for patients in the acute phase of injury as within-session ABCD indices remained relatively stable among patients at follow-up. Most control participants and patients at follow-up demonstrated ‘D’-type dynamics across the majority of EEG channels and blocks, which suggests that individuals with globally preserved thalamocortical network integrity may be less prone to state fluctuations than those with disrupted networks. As such, the within-session temporal variability we detected in acute patients may be a consequence of a more vulnerable thalamocortical network incapable of sustaining a consistently high level of functioning. Alternatively, this result may have been influenced by concomitant medical causes of altered consciousness (e.g., renal or hepatic dysfunction) or sedation. The majority (8/12) of follow-up patients showed recovery from PTCS. Thus, with only four patients who did not recover, we lacked statistical power to determine whether patients with chronic DoCs experience similar within-session temporal variability to that observed in patients with acute DoCs.

Unexpectedly, some healthy control participants demonstrated inter-block fluctuations between ‘C’- and ‘D’-type dynamics. Of note, we instructed all participants to keep their eyes closed to maintain consistency of EEG recording characteristics with patients incapable of eye-opening. Furthermore, participants were not disturbed by members of the investigational team during the 2-hour battery of EEG paradigms with the exception of administration of the CRS-R arousal facilitation protocol between blocks four and five. To maintain consistency across participants, we did not screen or reject any data based on the presence of EEG features suggestive of sleep given the lack of reliable electrophysiologic markers of sleep in acute brain injury.^43^ It is therefore possible that some control participants became drowsy or fell asleep during a portion of the 2-hour battery of EEG paradigms, resulting in EEG spectra with peaks in the theta frequency range and classification into the ‘C’ category. It is notable, however, that without exception, all controls demonstrated ‘D’-type signals across all channels during the first rest block and immediately following administration of the CRS-R arousal facilitation protocol (Fig. 2B). By contrast, patients demonstrated a range of ABCD indices during the same blocks, which further suggests that the state fluctuations observed in healthy participants may be attributable to sleep or drowsiness.

### ABCD classification as a potential biomarker for recovery of consciousness

As a group, longitudinally studied patients demonstrated an improvement in EEG signal dynamics at ≥6-month follow-up as compared to the acute setting. The three follow-up patients who were diagnosed as VS or MCS-had lower ABCD indices and demonstrated predominantly ‘B’- and ‘C’-type dynamics at follow-up. By contrast, the nine patients who recovered behavioural command-following at follow-up (MCS+ or R-PTCS) had significantly higher ABCD indices and predominantly demonstrated ‘D’-type dynamics (Figs. 3C, 4A). This separation in EEG dynamics at the level of MCS+/- underscores the crucial role of language function in recovery from severe brain injury and supports the notion that the pathophysiologic state of MCS-may be closer to VS rather than MCS+.^55^ Furthermore, the stark difference in ABCD indices between these two groups suggests that ABCD classification may prove to be a useful biomarker for recovery of consciousness. Specifically, it has potential to aid in early prognostication, provide a metric for monitoring recovery, and help identify individuals who should undergo additional testing with task-based paradigms to search for CMD.

Emerging evidence indicates that individuals with CMD constitute an estimated 15-20% of behaviourally unresponsive patients with a DoC.^17,20,24,56-59^ There is an ethical imperative to identify these individuals^60-62^ and ABCD classification may serve as an inexpensive, safe, and mobile screening measure well-suited to this purpose, especially when used as an adjunct to other similarly convenient resting-state EEG tools.^63^

We did not detect a similar separation in the ABCD indices of acute patients with and without behavioural command-following. Interestingly, however, one acute patient (P4) was behaviourally diagnosed as PTCS but demonstrated ‘A’-type dynamics in the majority of EEG channels (mean 56.7%, Figs. 1, 4B). It is possible that this discrepancy, as well as our inability to resolve a difference in ABCD indices among acute patients with and without command-following, are attributable to the rapid state fluctuations that we observed in the acute cohort (Fig. 2). However, this individual patient also demonstrated ‘D’-type dynamics in a mean of 28% of channels, and the coexistence of widespread cortical injury and partial thalamocortical network preservation has been previously observed in covertly conscious patients.^64,65^ This phenomenon warrants further study, but our findings suggest that only a limited amount of thalamocortical functional connectivity is necessary for the generation of consciousness. Therefore, channel-level analysis of EEG signal dynamics may be preferable to approaches that treat the entire cortex as a single entity,^7,36^ especially in the setting of multifocal cerebral injury.

It is also notable that a wide range of ABCD indices were demonstrated by acutely comatose patients. One of the two coma patients with predominantly ‘D’-type dynamics acutely went on to recover to R-PTCS, while the other recovered to MCS- and demonstrated a worsening of EEG signal dynamics at follow-up. By contrast, one of the three coma patients with predominantly ‘B’- or ‘C’- type dynamics acutely recovered to R-PTCS, while the other two recovered to MCS- and VS. Our observations suggest a need for larger studies to assess the prognostic relevance of the ABCD framework for the acute severe TBI population. The wide variability among acutely comatose patients also highlights the ethical obligation to further investigate patients who exhibit a dissociation between neurophysiologic measurements and behaviour.^60-62,66,67^

### CRS-R arousal facilitation protocol as a causal test of the mesocircuit model

To our knowledge, our observations serve as the first demonstration that the CRS-R arousal facilitation protocol effectively upregulates arousal in patients with DoCs to the point that it is detectable with neurophysiologic measurements (Fig. 5). Administration of the CRS-R arousal facilitation protocol involves sustained, rhythmic application of deep pressure stimulation to the face, sternocleidomastoid, trapezius, arm, and leg muscles,^39^ which produces a nonspecific afferent input into the anterolateral system (Fig. 6). Canonically, ascending anterolateral neurons innervate the ventral posterolateral and ventromedial nuclei of the thalamus, but these ascending neurons also innervate the lateral wing of the central lateral nucleus of the thalamus.^68^ Increasing evidence identifies the central lateral nucleus as a thalamic hub for the consciousness-supporting thalamocortical network, serving as the link between the anterior forebrain mesocircuit and the frontoparietal network.^30,69-71^ Given that the ABCD framework is theorized to index the functioning of the thalamocortical network, it follows that administration of the arousal facilitation protocol would yield improved EEG signal dynamics in this context through activation of thalamocortical afferents emanating from the central lateral nucleus.

**Figure 6.**
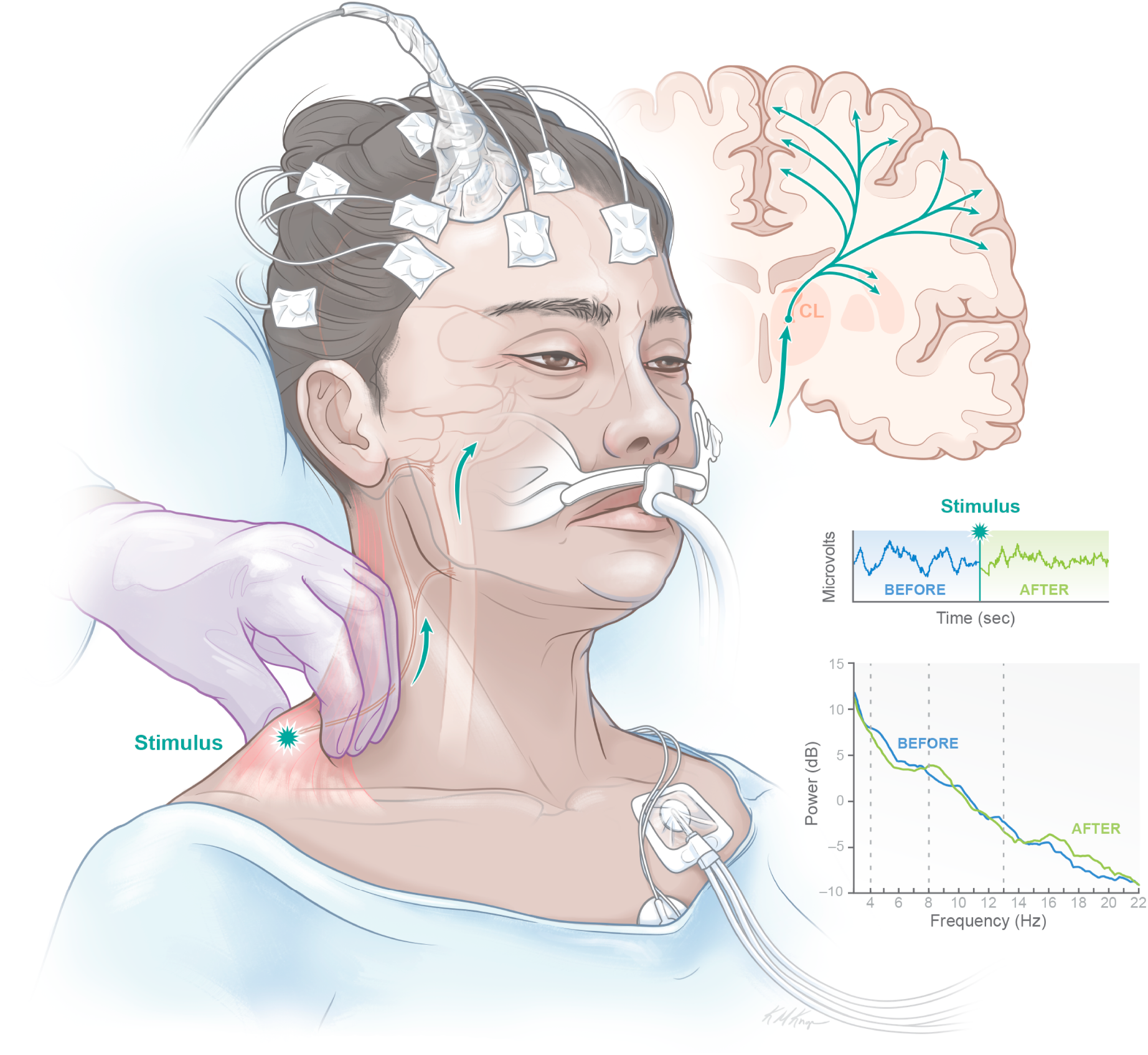
Schematic of proposed CRS-R arousal facilitation protocol mechanism. Administration of deep pressure muscle stimulation according to the CRS-R arousal facilitation protocol (stimulus) produces a nonspecific afferent input into the anterolateral system (green arrows), which projects to the lateral wing of the central lateral nucleus (orange; CL) in the thalamus. The central lateral nucleus in turn projects to striatum and cortex, thereby increasing excitatory neurotransmission throughout the cortex and shifting electrophysiologic dynamics towards a ‘D’-type pattern on the ABCD scale. EEG tracings and power spectra are shown from a representative patient studied acutely (P11). Vertical dotted lines on power spectra indicate separation of delta (<4 Hz), theta (4-8 Hz), alpha (8-13 Hz), and beta (13-24 Hz) frequency ranges. Artwork by Kimberly Main Knoper. dB = decibels, Hz = Hertz.

Furthermore, our analysis serves as a causal test that the mesocircuit model, and its instantiation as the ABCD framework, captures expected changes in functional thalamocortical network integrity. According to the model, increased functional integrity of the thalamocortical network produced by the arousal facilitation protocol results from the direct driving of the central thalamus, which in turn produces increased excitatory tone across the neocortex reflected in measured EEG dynamics (Fig. 6). If thalamocortical connectivity remains intact, the model predicts that dynamics should consistently shift in the direction of ‘D’ on the ABCD scale, whereas if structural injuries to the thalamocortical network are too severe, such changes in both ABCD dynamics and arousal should not occur. Studies applying the ABCD framework to smaller samples of patients have examined other gradations in functional and structural integrity of the thalamocortical network, including a predicted anterior-posterior gradient in ‘B,’ ‘C,’ and ‘D’ patterns linked to variations in drug-induced arousal state (and, in one patient, variations in cerebral resting metabolism).^31^ The pathological linkage of theta and beta/gamma oscillations in an isolable dynamical component of the resting EEG generated along the edge of cortical lesions has also been established.^72^ Our observation that stimulation of the central thalamus via the CRS-R arousal facilitation protocol yields improved ABCD signal dynamics provides unique supporting evidence that the model is indexing thalamocortical network function. Nonetheless, further study is needed to more firmly establish pathophysiologic linkages.

### Limitations and future directions

Our method for classifying EEG signals into ABCD categories relied on visual inspection of power spectral estimates. While this method has been demonstrated to have high inter-rater reliability,^7^ it is labor-intensive and requires expertise in EEG spectral analysis. Development of an automated method for ABCD classification is a key goal for future studies, as it would allow for efficient analysis of large-scale datasets and EEG assessments by clinicians without spectral analysis experience.

The acute EEG and behavioural data for the four patients enrolled at ≥6-months post-injury (P17-20) were collected under different conditions than the 16 prospectively enrolled patients (P1-16), in that they did not have discrete rest blocks interspersed within a larger study protocol in the ICU^23^ and behavioural level of consciousness was derived from clinical neurological exams instead of the CRS-R. As a result, there were likely inherent differences in the study environment between these two groups because for the former group, investigators were not present at the bedside to ensure minimization of distractors and to conduct other components of the study protocol (e.g., CRS-R arousal facilitation protocol, etc.). Additionally, lack of CRS-R assessments for some patients in the acute setting may have led to underestimation of level of consciousness.

We were also limited by sample size in this study, as we were only able to study 12 patients at follow-up. As such, we were not able to investigate the prognostic relevance of ABCD classification as it pertains to the acute severe TBI population. Recovery from severe brain injury is known to be variable in both time course and trajectory, and chronic CMD can emerge late in recovery.^17,64,66^ Therefore, larger studies with long-term follow-up are needed to assess the prognostic relevance of ABCD classification in TBI and shed light on the optimal application of the approach (i.e., whether cross-channel averaging to calculate an ABCD index is preferable to considering the relative proportions of each ABCD category). Results from a prior study were suggestive of a potential prognostic utility of ABCD classification in a mixed sample of acute non-hypoxic-ischemic patients (7% TBI),^36^ but additional studies investigating the characteristics of ABCD-classified EEG signals in other etiologies of severe brain injury are needed to determine the generalizability of this approach.

To minimize bias and maintain methodologic consistency across participants, we did not screen or reject EEG data on the basis of sleep features. We also did not investigate the potential effect of sedatives on our results because of the many different medications administered that do not have well-established equivalencies and because standardized sedation rating scales may not apply to patients with severe TBI.^73,74^ As such, it is possible that some results may have been influenced by sleep or, for patients studied acutely, sedation. However, ABCD classification is not designed to be used in isolation, especially in the acute setting. As with any method for evaluating patients with severe brain injuries, it would potentially be used as part of a multimodal assessment of brain function that takes into account tests of metabolic and hemodynamic function, in conjunction with behavioural, neuroimaging, and other EEG-based measures.^53,54,75^

### Conclusions

Based on the mesocircuit model for DoCs, resting-state EEG power spectra may be classified into categories (‘A,’ ‘B,’ ‘C,’ ‘D’) corresponding to graded levels of thalamocortical network function. Application of this approach to a cohort of patients with acute severe TBI allowed for identification of improvement in thalamocortical network function among patients followed up at ≥6-months post-injury as well as improved network functioning among patients who recovered behavioural command-following. Furthermore, ABCD classification captured rapid state fluctuations among acutely injured patients as well as neurophysiologic evidence for the effectiveness of the CRS-R arousal facilitation protocol. If validated in future studies, ABCD classification of EEG may emerge as a useful complement to task- and stimulus-based techniques already endorsed for use in the clinical setting for diagnosis and prognosis of patients with DoCs.^53,54^

## Acknowledgements

The authors thank the patients that participated in this study, as well as their families. We also thank the EEG technologists who facilitated data collection and helped make this study possible. We thank Kimberly Main Knoper for production of the artwork in Figure 6.

## Funding

This work was supported by the NIH National Institute of Neurological Disorders and Stroke (R21NS109627, RF1NS115268), NIH Director’s Office (DP2HD101400), James S. McDonnell Foundation, Tiny Blue Dot Foundation, National Institute on Disability, Independent Living and Rehabilitation Research (NIDILRR), Administration for Community Living (90DP0039, Spaulding-Harvard TBI Model System), Harvard Medical School Office of Scholarly Engagement, and NIH Shared Instrument Grant S10RR023043.

## Competing interests

Dr. Giacino occasionally receives honoraria for conducting CRS-R training seminars. The other authors report no competing interests.

### Abbreviations

CAP: Confusional Assessment Protocol
CI: confidence interval
CMD: cognitive motor dissociation
CRS-R: Coma Recovery Scale – Revised
dB: decibels
DoC: disorder of consciousness
EMCS: emerged from minimally conscious state
functional MRI: fMRI
GCS: Glasgow Coma Scale
Hz: Hertz
ICU: intensive care unit
IQR: interquartile range
MCS+/-: minimally conscious state with/without language
PTCS: post-traumatic confusional state
RMSD: root mean square deviation
R-PTCS: recovered from PTCS
SD: standard deviation
TBI: traumatic brain injury
VS: vegetative state

## Notes

### Funding Statement

This work was supported by the NIH National Institute of Neurological Disorders and Stroke (R21NS109627, RF1NS115268), NIH Office of the Director (DP2HD101400), James S. McDonnell Foundation, Tiny Blue Dot Foundation, National Institute on Disability, Independent Living and Rehabilitation Research (NIDILRR), Administration for Community Living (90DP0039, Spaulding-Harvard TBI Model System), Harvard Medical School Office of Scholarly Engagement, and NIH Shared Instrument Grant S10RR023043.

### Author Declarations

All studies were approved by the Mass General Brigham Institutional Review Board.

