## Supplementary Material for "Electrophysiological correlates of thalamocortical function in acute severe traumatic brain injury"

**Supplementary Table 1. Sedative, anxiolytic, and analgesic medications administered before and/or during EEG.**

| Patient ID | Medications Administered Before/During EEG |
|---|---|
| P1 | **Before:** None<br>**During:** None |
| P2 | **Before:** morphine 2 mg IV<br>**During:** None |
| P3 | **Before:** propofol 100 mg/hr IV gtt<br>**During:** propofol 100 mg/hr gtt, propofol 10 mg IV bolus |
| P4 | **Before:** None<br>**During:** None |
| P5 | **Before:** oxycodone 5 mg PGT<br>**During:** quetiapine 25 mg PGT |
| P6 | **Before:** hydromorphone 0.5 mg IV<br>**During:** None |
| P7 | **Before:** quetiapine 12.5 mg PGT<br>**During:** None |
| P8 | **Before:** propofol 300 mg/hr IV gtt<br>**During:** propofol 300 mg/hr IV gtt, hydromorphone 0.5 mg IV |
| P9 | **Before:** None<br>**During:** None |
| P10 | **Before:** None<br>**During:** None |
| P11 | **Before:** lorazepam 1 mg IV, haloperidol 5 mg IV<br>**During:** None |
| P12 | **Before:** midazolam 2 mg IV<br>**During:** None |
| P13 | **Before:** None<br>**During:** None |
| P14 | **Before:** None<br>**During:** None |
| P15 | **Before:** propofol 300 mg/hr IV gtt, fentanyl 50 mcg IV bolus<br>**During:** propofol 300 mg/hr IV gtt |
| P16 | **Before:** None<br>**During:** None |
| P17 | [a]**Before/During:** None |
| P18 | [a]**Before/During:** propofol 250 mg/hr IV gtt, oxycodone 15 mg PGT, fentanyl 100 mcg IV bolus |
| P19 | [a]**Before/During:** propofol 100 mg/hr IV gtt, fentanyl 50 mcg/hr IV gtt, fentanyl 100 mcg IV bolus |
| P20 | [a]**Before/During:** propofol 10 mg/hr IV gtt, oxycodone 10 mg PGT |

[a]As patients P17-20 were enrolled at follow-up, the precise timing of medication administration during the acute phase could not be definitively determined. As such, we list all possible medications these patients may have received before and/or during acute EEG collection.
gtt = continuous infusion; IV = intravenous; PGT = administered via percutaneous gastric tube.

**STROBE statement: Reporting guidelines checklist for cohort, case-control and cross-sectional studies.**

| SECTION | ITEM NUMBER | CHECKLIST ITEM | REPORTED ON PAGE NUMBER: |
|---|---|---|---|
| **TITLE AND ABSTRACT** | | | |
| | 1a | Indicate the study's design with a commonly used term in the title or the abstract | 2 |
| | 1b | Provide in the abstract an informative and balanced summary of what was done and what was found | 2 |
| **INTRODUCTION** | | | |
| Background and objectives | 2 | Explain the scientific background and rationale for the investigation being reported | 3-4 |
| | 3 | State specific objectives, including any pre-specified hypotheses | 4 |
| **METHODS** | | | |
| Study design | 4 | Present key elements of study design early in the paper | 4-8 |
| Setting | 5 | Describe the setting, locations, and relevant dates, including periods of recruitment, exposure, follow-up, and data collection | 4 |
| Participants | 6a | Cohort study—Give the eligibility criteria, and the sources and methods of selection of participants. Describe methods of follow-up<br>Case-control study—Give the eligibility criteria, and the sources and methods of case ascertainment and control selection. Give the rationale for the choice of cases and controls<br>Cross-sectional study—Give the eligibility criteria, and the sources and methods of selection of participants | 4 |
| | 6b | Cohort study—For matched studies, give matching criteria and number of exposed and unexposed<br>Case-control study—For matched studies, give matching criteria and the number of controls per case | N/A |
| Variables | 7 | Clearly define all outcomes, exposures, predictors, potential confounders, and effect modifiers. Give diagnostic criteria, if applicable | 5-8 |
| Data sources/measurements | 8* | For each variable of interest, give sources of data and details of methods of assessment (measurement). Describe comparability of assessment methods if there is more than one group. | 4-8 |
| Bias | 9 | Describe any efforts to address potential sources of bias. | 4-8 |
| Study size | 10 | Explain how the study size was arrived at | 4 |
| Quantitative variables | 11 | Explain how quantitative variables were handled in the analyses. If applicable, describe which groupings were chosen and why . | 4-8 |
| Statistical methods | 12a | Describe all statistical methods, including those used to control for confounding | 7-8 |
| | 12b | Describe any methods used to examine subgroups and interactions | 4-8 |
| | 12c | Explain how missing data were addressed | 4-8 |
| | 12d | Cohort study—If applicable, explain how loss to follow-up was addressed<br>Case-control study—If applicable, explain how matching of cases and controls was addressed<br>Cross-sectional study—If applicable, describe analytical methods taking account of sampling strategy | 4 |
| | 12e | Describe any sensitivity analyses | N/A |
| **RESULTS** | | | |
| Participants | 13a | Report numbers of individuals at each stage of study—eg numbers potentially eligible, examined for eligibility, confirmed eligible, included in the study, completing follow-up, and analysed | 8 |
| | 13b | Give reasons for non-participation at each stage | 5,8 |
| | 13c | Consider use of a flow diagram | N/A |

| SECTION | ITEM NUMBER | CHECKLIST ITEM | REPORTED ON PAGE NUMBER: |
|---|---|---|---|
| Descriptive Data | 14a | Give characteristics of study participants (eg demographic, clinical, social) and information on exposures and potential confounders | 8 |
| | 14b | Indicate number of participants with missing data for each variable of interest | 8-12 |
| | 14c | Cohort study—Summarise follow-up time (eg, average and total amount) | 8 |
| Outcome Data | 15* | Cohort study—Report numbers of outcome events or summary measures over time<br>Case-control study—Report numbers in each exposure category, or summary measures of exposure<br>Cross-sectional study—Report numbers of outcome events or summary measures | N/A |
| Main Results | 16a | Give unadjusted estimates and, if applicable, confounder-adjusted estimates and their precision (e.g. 95% confidence interval). Make clear which confounders were adjusted for and why they were included | 8-12 |
| | 16b | Report category boundaries when continuous variables were categorized | 8-12 |
| | 16c | If relevant, consider translating estimates of relative risk into absolute risk for a meaningful time period | N/A |
| | 16d | Report results of any adjustments for multiple comparisons | N/A |
| Other Analyses | 17a | Report other analyses done—e.g. analyses of subgroups and interactions, and sensitivity analyses | 9-12 |
| | 17b | If numerous genetic exposures (genetic variants) were examined, summarize results from all analyses undertaken | N/A |
| | 17c | If detailed results are available elsewhere, state how they can be accessed | N/A |
| **DISCUSSION** | | | |
| Key Results | 18 | Summarise key results with reference to study objectives | 13 |
| Limitations | 19 | Discuss limitations of the study, taking into account sources of potential bias or imprecision. Discuss both direction and magnitude of any potential bias | 14-15,17 |
| Interpretation | 20 | Give a cautious overall interpretation of results considering objectives, limitations, multiplicity of analyses, results from similar studies, and other relevant evidence | 13-18 |
| Generalisability | 21 | Discuss the generalisability (external validity) of the study results | 14-18 |
| **FUNDING** | | | |
| | 22 | Give the source of funding and the role of the funders for the present study and, if applicable, for the original study on which the present article is based | 18 |
